# Patients’ and Clinicians’ Perceptions of Motivational Factors in Rehabilitation

**DOI:** 10.1101/2022.09.30.22280550

**Authors:** Kazuaki Oyake, Katsuya Yamauchi, Seigo Inoue, Keita Sue, Hironobu Ota, Junichi Ikuta, Toshiki Ema, Tomohiko Ochiai, Makoto Hasui, Yuya Hirata, Ayaka Hida, Kenta Yamamoto, Yoshihiro Kawai, Kiyoto Shiba, Akihito Atsumi, Tetsuyuki Nagafusa, Satoshi Tanaka

## Abstract

**Importance:** Patient motivation is an important determinant of rehabilitation outcomes. Differences in patients’ and clinicians’ perceptions of motivational factors can potentially hinder patient-centered care.

**Objective:** To compare patients’ and clinicians’ perceptions of the most important factors in motivating patients for rehabilitation.

**Design:** This multicenter descriptive cross-sectional survey was conducted from January to March 2022.

**Setting:** Thirteen hospitals with a convalescent rehabilitation ward.

**Participants:** Patients with neurological or orthopedic disorders undergoing inpatient rehabilitation and clinicians, including physicians, physical therapists, occupational therapists, and speech-language-hearing therapists, were selected purposively based on the inclusion criteria.

**Main Outcomes and Measures:** Patients and clinicians were asked to choose the most important factor from a list of potential motivational factors. The main outcome was patients’ and clinicians’ perceptions of the relative importance of various motivational factors for rehabilitation.

**Results:** We obtained data from 479 patients and 401 clinicians. Response rates in the patient and clinician surveys were 92.1% and 62.2%, respectively. The most common primary reasons for patients’ hospitalizations were stroke (45.5%) and fracture (42.2%). Approximately half of the clinicians were physical therapists (49.9%). “Realization of recovery,” “goal setting,” and “practice related to the patient’s experience and lifestyle” were the three factors most frequently selected as most important by both patients and clinicians, chosen by 10.4%–26.5% of patients and 9.5%–36.7% of clinicians. Only five were rated as most important by 5% of clinicians; however, nine factors were selected by 5% of patients. Of these nine motivational factors, “medical information” (odds ratio: 5.19; 95% confidence interval: 2.24– 11.60) and “control of task difficulty” (odds ratio: 2.70; 95% confidence interval: 1.32–5.80) were selected by a significantly higher proportion of patients than clinicians.

**Conclusions and Relevance:** The three most frequently endorsed motivational factors were identical for patients and clinicians. The preferences of patients were more diverse than those of clinicians, and some motivational factors were preferred by patients over clinicians. Therefore, when determining motivational strategies, rehabilitation clinicians should consider individual patient preferences in addition to utilizing the core motivational factors supported by both parties.

**Key Points:** *Question:* What are the similarities and differences between patients’ and clinicians’ perceptions of the relative importance of factors motivating patients for rehabilitation?

*Findings:* In this multicenter descriptive cross-sectional survey of 479 patients and 401 clinicians, the three most endorsed motivational factors—realization of recovery goal setting, and practice related to the patient’s experience and lifestyle—were identical for patients and clinicians. However, patients had more diverse preferences for motivational factors than clinicians.

*Meaning:* In addition to utilizing the three core motivational factors, rehabilitation clinicians should consider individual patient preferences when determining which motivational strategies to use for enhancing patient-centered care.

## INTRODUCTION

For patients with physical disabilities, rehabilitation programs including physical activity and exercise have beneficial effects on several health outcomes, such as improved physical function and enhanced health-related quality of life.^1^ However, physical activity levels are lower among patients with physical disabilities than among those without disabilities.^1-3^ As a lack of motivation is often the main barrier to physical activity and exercise training,^4-10^ motivation is frequently considered to be a determinant of rehabilitation outcomes.^11-15^ Thus, various strategies, such as motivational interviewing and behavioral therapies, are used to enhance patient motivation for rehabilitation with positive motor and functional outcomes for individuals with a range of neurological and orthopedic disorders.^16-26^ Motivation has been defined as the “mental function that produces the incentive to act; the conscious or unconscious driving force for action.”^27^ Social factors, such as clinicians’ attitudes towards the patient and the patient’s social support networks, in combination with the personality or clinical characteristics of the individual patient have been considered possible determinants of motivation for rehabilitation.^4-10,28^ For example, the opportunities to meet other people who have suffered strokes and group exercise have been reported as perceived motivators of physical activity in individuals with strokes.^4^ Conversely, our previous Delphi study indicated that rehabilitation experts rated “group rehabilitation” as neither effective nor ineffective in motivating patients with strokes.^29^ Based on these findings, we hypothesized that patients and clinicians would to some extent differ in their perceptions of the relative importance of factors that motivate patients for rehabilitation. The differences in patients’ and clinicians’ perceptions of motivational factors could potentially hinder patient-centered care.^30^ Patient-centered care is a core principle of evidence-based medical practice and is more likely to positively affect rehabilitation outcomes, such as improvements in functional performance and quality of life.^31,32^ However, to the best of our knowledge, no studies have directly compared patients’ preferences regarding motivational factors with those of clinicians. Therefore, the current study aims to compare patients’ and clinicians’ perceptions of the most important factors in motivating patients for rehabilitation.

## METHODS

### Study design

We employed a multicenter descriptive cross-sectional design. This study protocol was approved by the appropriate ethics committee at the Hamamatsu University School of Medicine (approval number: 21-233). Informed consent was obtained from all participants.

### Participants

Patients hospitalized in a convalescent rehabilitation ward were recruited through purposive sampling based on the inclusion criteria from 12 hospitals in Japan. In convalescent rehabilitation wards, intensive rehabilitation programs are provided during hospitalization to improve inpatients’ abilities to perform activities of daily living.^33,34^ All rehabilitation inpatients meeting the inclusion criteria were referred to the research team by a researcher at each hospital. Inclusion criteria were as follows: being aged 20– 90 years, having an established diagnosis of neurological or orthopedic disorders as the primary reason for hospitalization, having undergone an inpatient rehabilitation program for at least four weeks at the time of study participation, and adequate communication skills to complete the questionnaire. Demographic and clinical data, such as primary reasons for hospitalization and sex, were obtained from patient medical records. Clinicians were also purposively sampled from 13 hospitals in Japan. They included physicians, physical therapists, occupational therapists, or speech-language-hearing therapists working in a convalescent rehabilitation ward. Patients were recruited as participants from 12 of these hospitals.

The sample size calculation for participants was based on epidemiological data from the Kaifukuki Rehabilitation Ward Association.^35^ Totally, 38,363 patients with neurological and orthopedic diseases were admitted to convalescent rehabilitation wards. Of them, 18,870 patients had neurological diseases and 19,493 had orthopedic diseases. Similarly, the total number of rehabilitation clinicians working in the convalescent rehabilitation wards was estimated at 66,033, with 30,911 physical therapists, 18,700 occupational therapists, 8,843 physicians, and 7,579 speech-language-hearing therapists. Based on these population sizes and using a margin of error of 5% at a 95% confidence interval (CI), the estimated minimum sample size was 381 patients and 382 clinicians for this study.^36^

### Questionnaire content

The first author initially developed a list of potential factors involved in increasing patient motivation for rehabilitation based on previous findings.^25,29,37^ Two researchers (K.S. and S.T.) reviewed the items for clarity, relevance, and topic coverage.^38^ We conducted a pilot test with a small sample of patients and clinicians to determine whether participants consistently understood the meaning of each item.^39^ Based on feedback from the pilot test, minor grammatical changes were made. Consequently, we prepared a list of 15 potential motivational factors for rehabilitation (Table 1). All survey data were captured anonymously for both patients and clinicians.

**Table 1.** List of potential factors in motivating patients for rehabilitation.

| Potential motivational factors | Specific examples |
| --- | --- |
| Active listening | Clinicians listen to patients carefully while showing interest, such as conducting motivational interviews to strengthen patients' sense of motivation and commitment to change. |
| A suitable rehabilitation environment | Clinicians provide a suitable rehabilitation environment so that patients can comfortably engage in rehabilitation practices. |
| Control of task difficulty | Clinicians gradually increase the difficulty of a task according to the ability of the patient. |
| Enjoyable rehabilitation programs | Clinicians support patients to enjoy themselves during the rehabilitation process by applying their preferences to the practice tasks, engaging in pleasant conversation with them, and providing a practice task with game-like properties. |
| Feedback regarding the results of the practice | Clinicians provide verbal and/or visual feedback to their patients regarding their progress and results of the rehabilitation practices. |
| Goal setting | Clinicians set rehabilitation goals for individual patients and provide practice tasks that are related to each patient's rehabilitation goal. |
| Group rehabilitation | Clinicians offer a group rehabilitation program. |
| Medical information | Patients receive medical information regarding topics such as the necessity of practice and the nature of recovery and what patients can expect in their lives at home. |
| Practice related to the patient's experience and lifestyle | Patients engage in practice tasks that they can complete using their previous experience. |
| Praise | Clinicians provide patients with positive evaluations and encouragement. |
| Presence of family members during rehabilitation | The patient's family members are present during rehabilitation. |
| Realization of recovery | Patients realize their own recovery, for example, (1) by using a diary or graph that helps to track their progress and (2) by acquiring new skills through rehabilitation practices. |
| Rehabilitation programs with variations | Clinicians provide variations of rehabilitation programs. |
| Respect for self-determination | Clinicians respect patients' self-determination and encourage them to consider how they could successfully perform the provided practice tasks. |
| Rewards for effort | Clinicians propose conditions for exchange to their patients. For example, clinicians promise that patients can do their favorite practice task after completing their least favorite practice task. |

Note. Potential motivational factors are arranged in alphabetical order.

We administered the patient questionnaire in an interview style, and patients took part in a face-to-face structured interview with a researcher at each hospital. Patients were presented with a list of 16 items, including “other” in addition to the 15 potential motivational factors. The structured interview included two questions. In the first question, patients were asked to select the three most important factors for facilitating their engagement in rehabilitation from the list. In the second question, they were instructed to choose the most important factor from the three factors they selected in the first question. Participants who selected “other” were asked to respond to an open-ended question in which they proposed additional motivational factors. The interview lasted less than five minutes. The patient survey was available in paper form. In each hospital, one researcher was responsible for administering the survey. Following the end of the recruitment period, all completed questionnaires were mailed to the first author.

We used a cloud-based questionnaire and survey software (Google Forms tool, Google LLC, Mountain View, CA, USA) to develop the clinician survey and to collect data. To publicize the study, the researcher at each hospital distributed leaflets to clinicians who met the inclusion criteria. The leaflets contained a brief description of the study and a hyperlink to the survey. Clinicians could voluntarily access the survey website using their own laptops, tablets, or smartphones. The survey included two questions and a few demographic characteristics. In the first question, clinicians were asked to select the three most important factors for increasing patient adherence to rehabilitation programs from the list. In the second question, they were also instructed to choose the most important factor out of the three they selected in the first question. Clinicians who completed the survey were reimbursed for their participation with a 500-yen gift card (approximately USD $3.50).

### Statistical analysis

The primary outcome of the study was patients’ and clinicians’ top choice for the most important motivational factor. The secondary outcome included the three factors that they selected in the first question. We used descriptive statistics to summarize the demographic characteristics of patients and clinicians and their responses to the two survey questions. Because we used a margin of error of 5% to determine the sample size of this study, motivational factors selected by more than 5% of participants in each of the patient and clinician groups were considered to constitute their preferences. We compared patients’ responses with those of clinicians by calculating odds ratios (ORs) with 95% CIs. When there was a null value in one of the four cells, we added 0.5 to all cells.^40^ In addition, a multiple logistic regression analysis was used to examine the association between patients’ choices regarding the most important motivational factor with their demographic characteristics, including the primary reason for hospitalization, sex, age ≥ 65 years or not,^41^ and the length of hospital stay. Statistical analyses were performed using the Statistical Package for the Social Sciences software version 27.0 (International Business Machines Corp., NY, USA).

## RESULTS

### Participants’ characteristics

The survey was conducted from January to March 2022. Of the total of 520 patients who met the inclusion criteria, 23 refused to participate in this study. Consequently, we obtained data from 479 patients. In addition, of the 645 clinicians who met the inclusion criteria, 401 responded. Thus, the response rates of the patient and clinician surveys were 92.1% and 62.2%, respectively. The demographic characteristics of patients and clinicians are presented in Table 2. The primary reason for hospitalization of most patients was either stroke (45.5%) or fracture (42.2%). Approximately half of the clinicians were physical therapists (49.9%).

**Table 2.** Characteristics of participants.

| Variable | Value |
| --- | --- |
| Patients (n = 479) |  |
| Primary reason for hospitalization |  |
| Stroke | 218 (45.5) |
| Fracture | 202 (42.2) |
| Spinal cord injury | 24 (5.0) |
| Amputation | 8 (1.7) |
| Arthritic disorders | 8 (1.7) |
| Traumatic brain injury | 8 (1.7) |
| Neuromuscular disorders | 5 (1.0) |
| Brain and spinal cord tumors | 4 (0.8) |
| Others | 2 (0.4) |
| Sex |  |
| Female | 266 (55.5) |
| Male | 213 (44.5) |
| Age | 79 [70–84] |
| Time since the onset of primary disease, days | 70.50 [57.00–97.25] |
| Length of hospital stay, days | 42.00 [33.00–63.00] |
| Clinicians (n = 401) |  |
| Professional category |  |
| Physical therapist | 200 (49.9) |
| Occupational therapist | 142 (35.4) |
| Speech-language-hearing therapist | 37 (9.2) |
| Physician | 22 (5.5) |
| Sex |  |
| Male | 214 (53.4) |
| Female | 187 (46.6) |
| Years of clinical experience | 5 [3–11] |

Values are presented as number (%) and median [interquartile range].

### Comparison of patients’ and clinicians’ perceptions of the most important factors for motivating patients to engage in rehabilitation

The distribution of patients’ and clinicians’ answers, when asked to report their top choice regarding the most important motivational factor, is shown in Figure 1. The three most frequently selected motivational factors were identical for patients and clinicians: “realization of recovery,” “goal setting,” and “practice related to the patient’s experience and lifestyle,” chosen by 10.4%–26.5% of patients and 9.5%–36.7% of clinicians.

**Figure 1.**
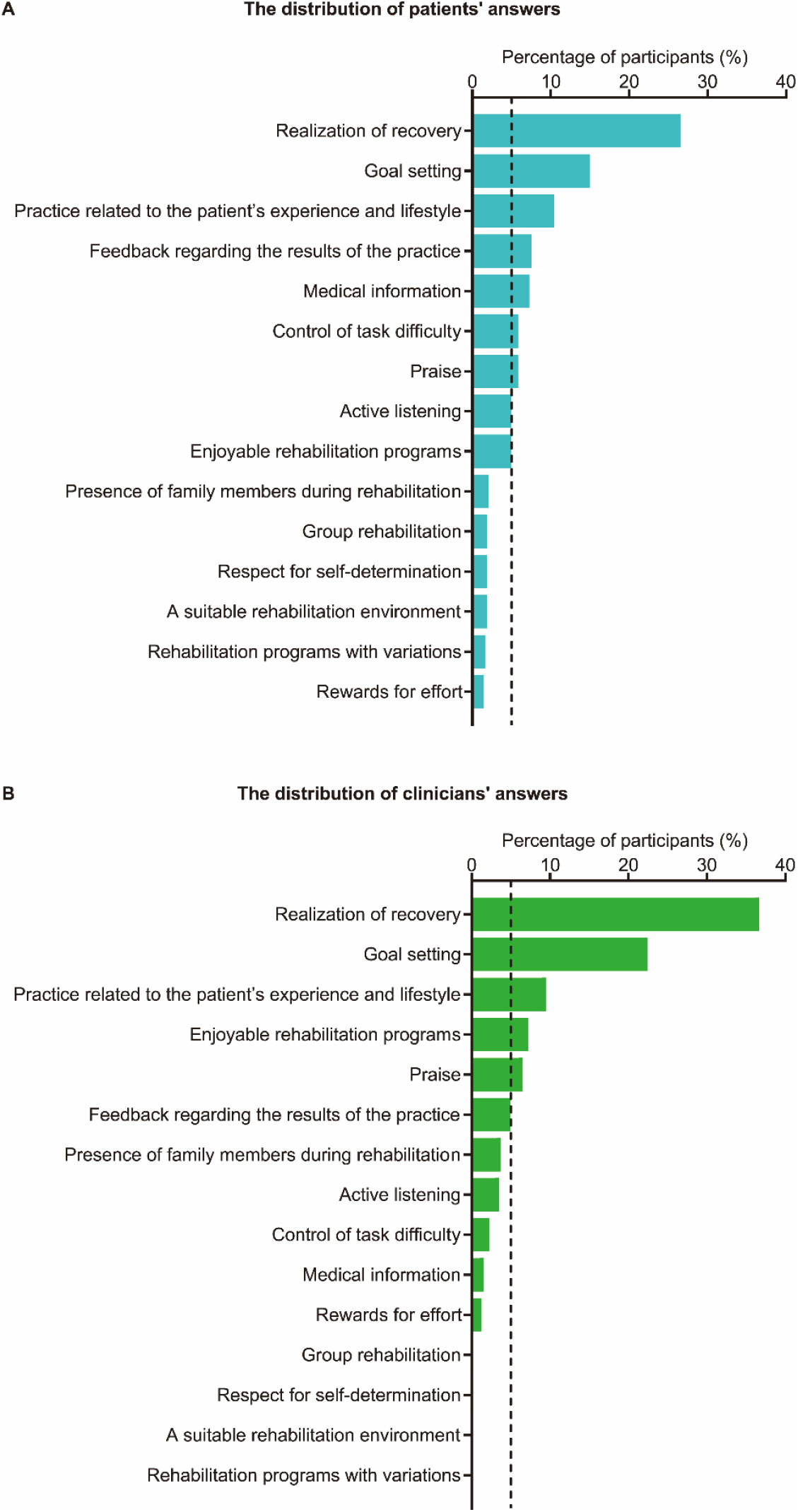
Distributions of patients’ (A) and clinicians’ (B) answers when asked to report their top choice for the most important motivational factor. Motivational factors are arranged in descending order by the percentage of participants who selected each factor as the most important. The vertical dashed line represents 5% of participants.

Although nine motivational factors were selected by more than 5% of the patients, only five were chosen by more than 5% of the clinicians, indicating that patients exhibited more varied preferences for motivational factors than clinicians (Figure 1). The ORs comparing patients’ top choices with those of clinicians are shown in Figure 2. Of the nine motivational factors selected by more than 5% of patients, “medical information” (OR: 5.19; 95% CI: 2.24–11.60) and “control of task difficulty” (OR: 2.70; 95% CI: 1.32–5.80) were chosen by a significantly higher proportion of patients than clinicians.

**Figure 2.**
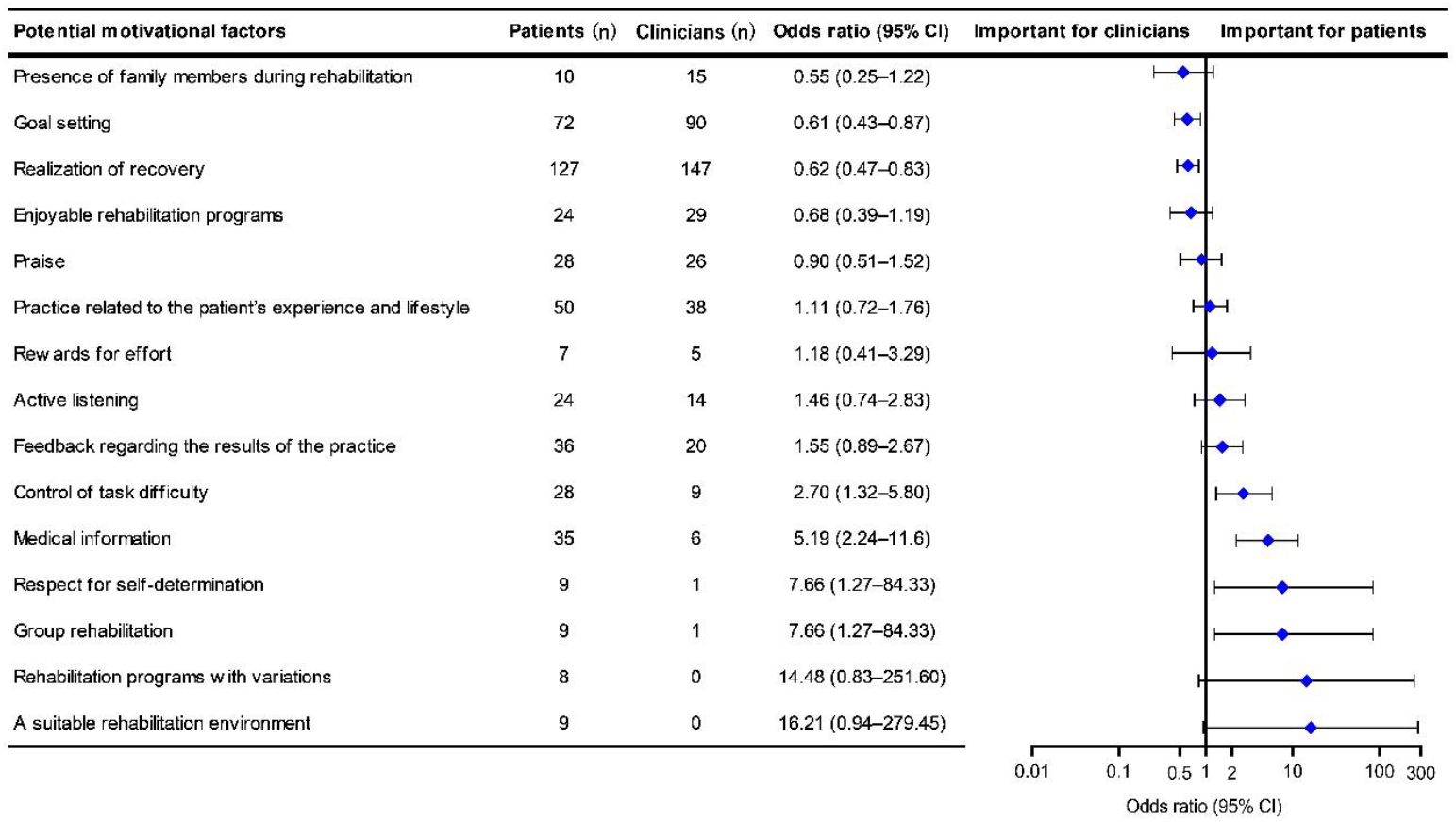
Comparison of patients’ and clinicians’ perceptions of the most important factors motivating patients for rehabilitation. Motivational factors are arranged in ascending order regarding the value of the odds ratio. Filled diamonds and horizontal bars represent the odds ratio and 95% confidence intervals, respectively.

### Comparison of patients’ and clinicians’ perceptions of the three most important motivational factors

The distribution of patients’ and clinicians’ answers, when asked to choose the three most important factors among 15 potential motivational factors, is shown in Figure 3. Twelve patients (2.5%) selected “other” as one of the three most important motivational factors. Additional motivational factors proposed by patients are shown in Table 3. Similar to the results on the motivational factors perceived as the most important, the three most frequently endorsed motivational factors were identical for patients and clinicians: “realization of recovery,” “goal setting,” and “practice related to the patient’s experience and lifestyle.” These factors were chosen by 32.4%– 47.4% of the patients and 38.2%–59.4% of the clinicians.

**Table 3.**
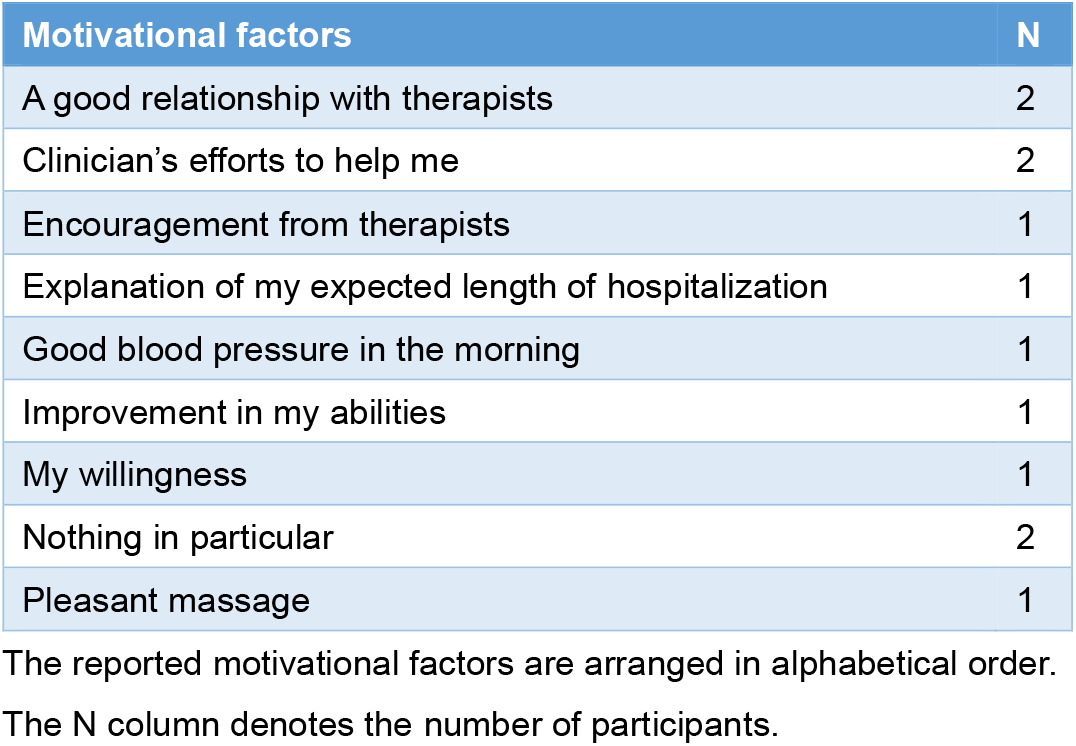
Additional factors motivating patients to engage in rehabilitation proposed by patients.

| Motivational factors | N |
| --- | --- |
| A good relationship with therapists | 2 |
| Clinician's efforts to help me | 2 |
| Encouragement from therapists | 1 |
| Explanation of my expected length of hospitalization | 1 |
| Good blood pressure in the morning | 1 |
| Improvement in my abilities | 1 |
| My willingness | 1 |
| Nothing in particular | 2 |
| Pleasant massage | 1 |

The reported motivational factors are arranged in alphabetical order.
The N column denotes the number of participants.

**Figure 3.**
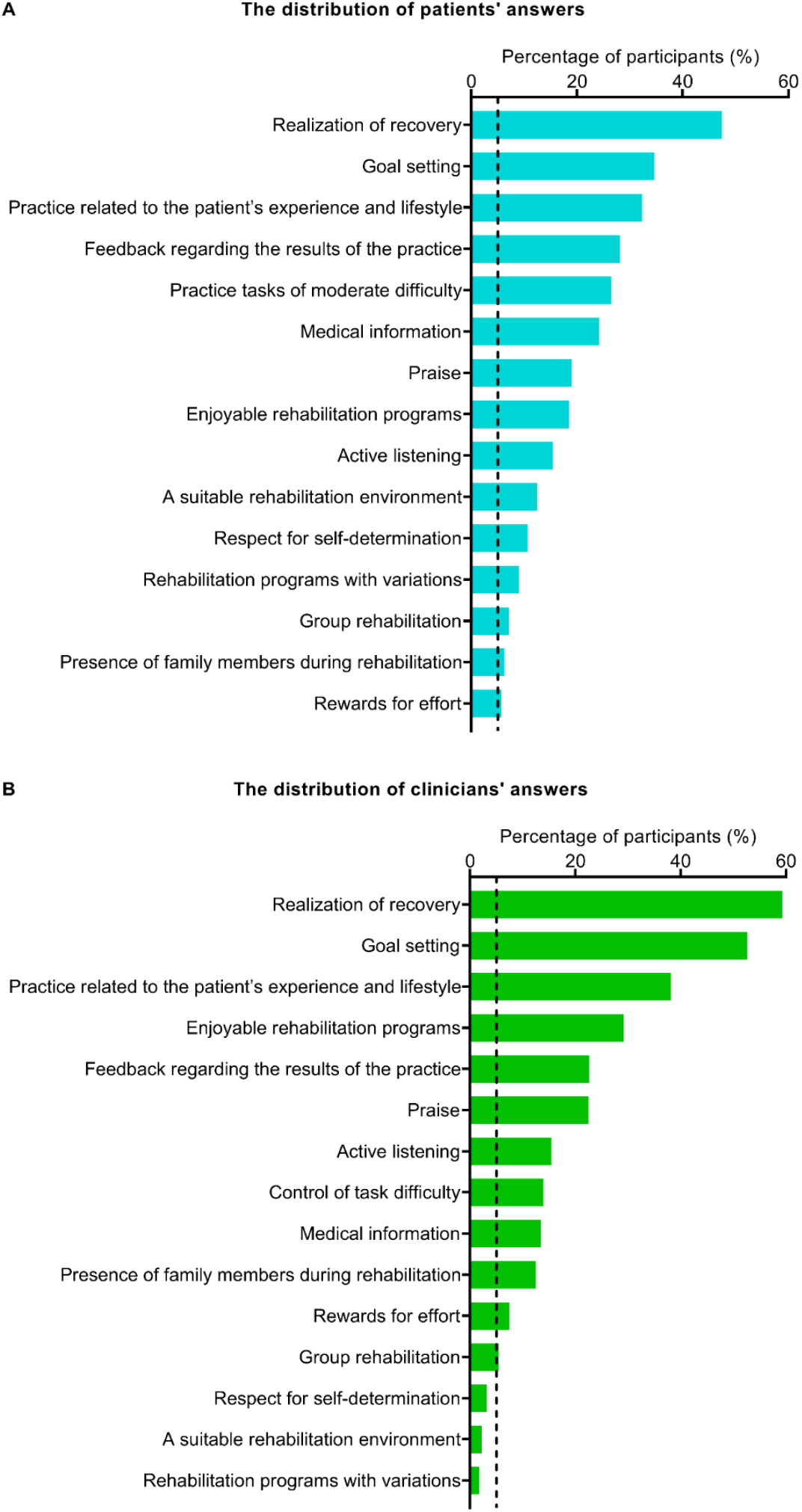
Distribution of patients’ (A) and clinicians’ (B) answers when asked to choose the three most important items among 15 potential motivational factors. Motivational factors are arranged in descending order by the percentage of patients who selected each item as one of the three most important factors. The vertical dashed line represents 5% of participants.

The ORs comparing patients’ choices with those of clinicians for the first question of the survey are shown in Figure 4. A significantly higher proportion of patients than clinicians rated “a suitable rehabilitation environment” (OR: 6.24; 95% CI: 3.14–12.42), “rehabilitation programs with variations” (OR: 5.55; 95% CI: 2.51–12.77), “respect for self-determination” (OR: 3.56; 95% CI: 1.93–6.58), “control of task difficulty” (OR: 2.22; 95% CI: 1.57–3.15), and “medical information” (OR: 2.05; 95% CI: 1.45–2.95), as important.

**Figure 4.**
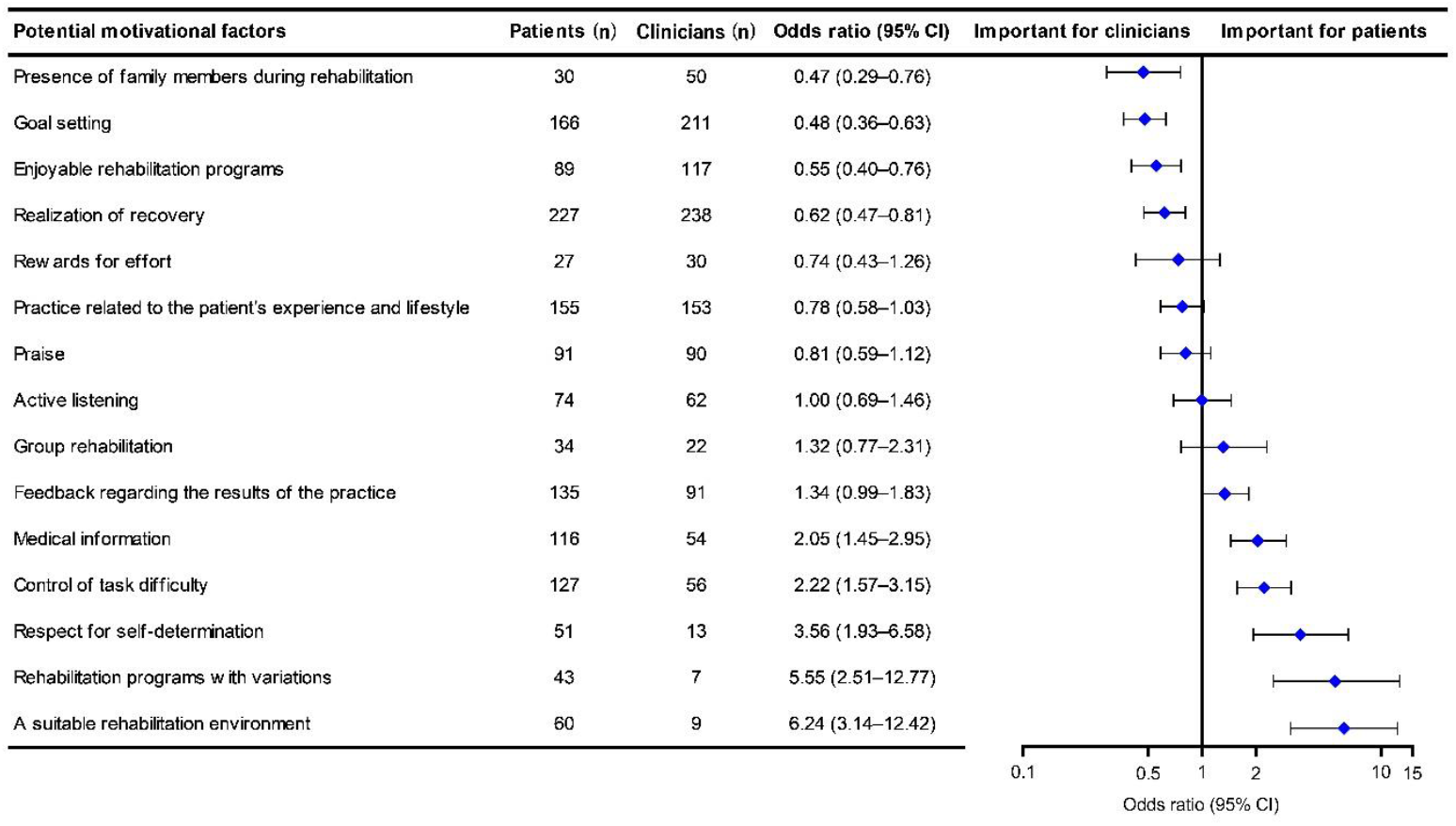
Comparison of patients’ and clinicians’ perceptions of the three most important factors motivating patients for rehabilitation. Motivational factors are arranged in ascending order regarding the value of the odds ratio. Filled diamonds and horizontal bars represent the odds ratio and 95% confidence intervals, respectively.

### Associations between patients’ choices regarding the most important motivational factors and their demographic characteristics

In patients with stroke and those with fracture (n = 420), who made up the majority of patient participants (87.7%), we evaluated the associations between patients’ choices regarding the most important motivational factor and their demographic characteristics. Most patients were those with stroke (51.9%), female (57.1%), and aged ≥ 65 years (84.3%). The median length of hospital stay was 43 days (interquartile range, 34– 64 days). The results of the multiple logistic regression analysis are shown in Table 4. A significantly higher proportion of patients aged < 65 years old, compared with those aged ≥ 65 years old, rated “goal setting” (OR: 0.46; 95% CI: 0.22– 0.93) and “realization of recovery” (OR: 0.53; 95% CI: 0.29–0.97) as the most important factor. Additionally, patients with shorter hospital stay lengths were significantly more likely to choose “medical information” as the most important factor (OR: 0.97; 95% CI: 0.94–1.00).

**Table 4.**
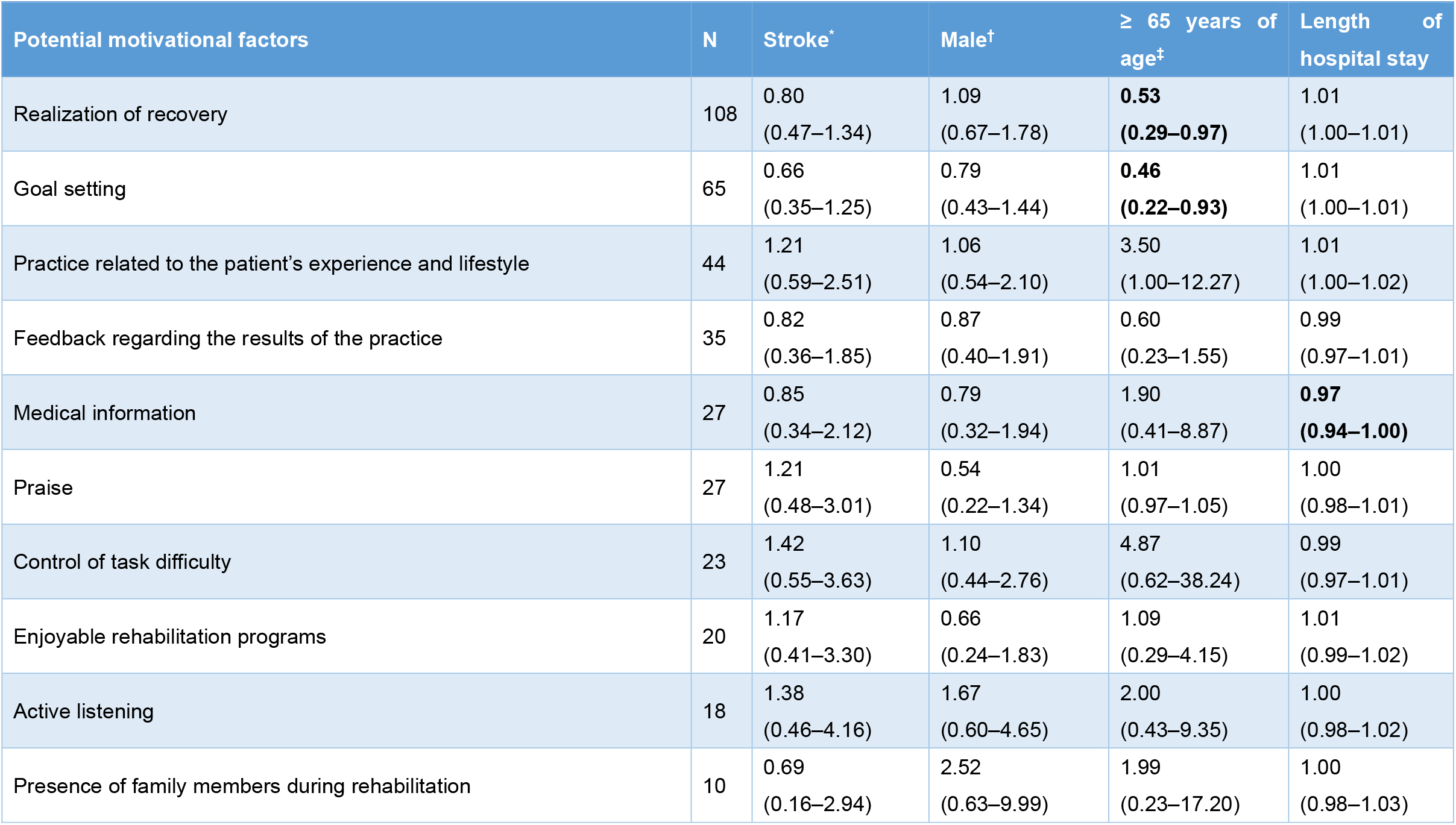
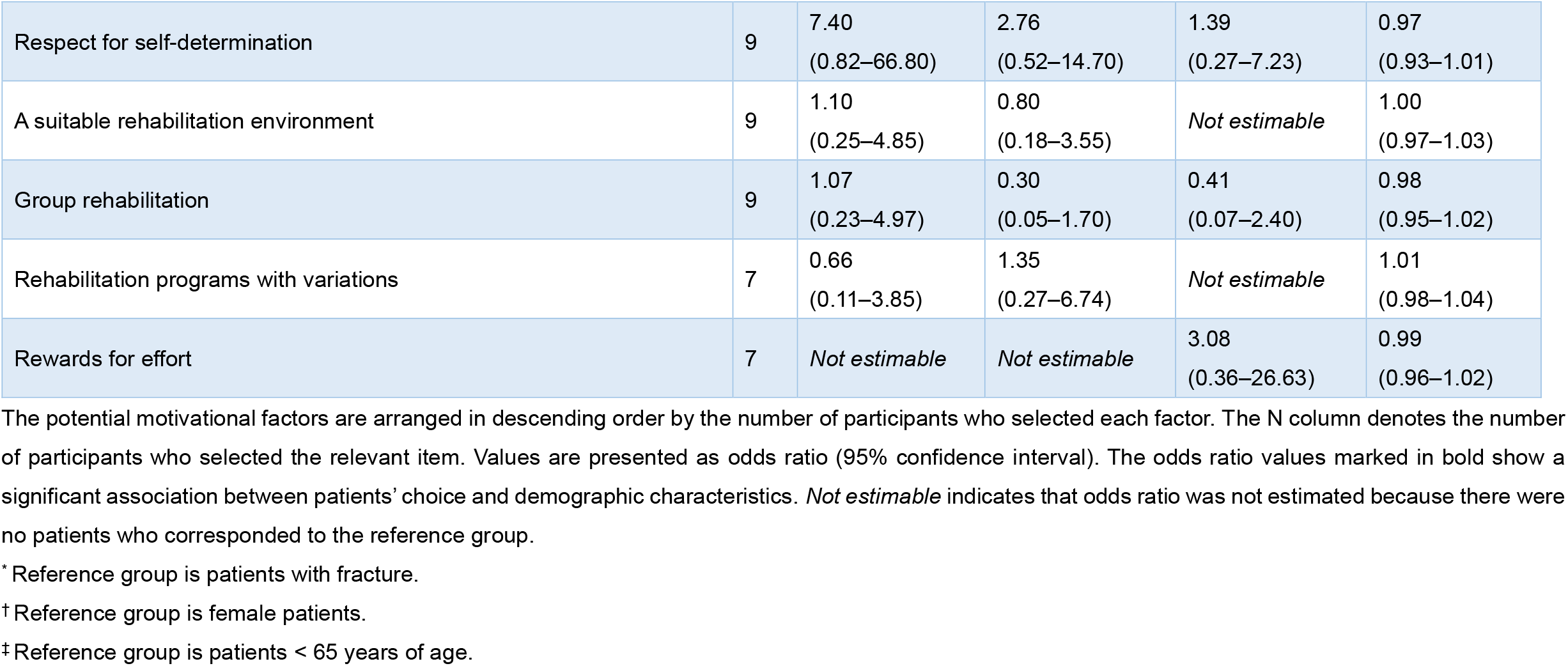
Associations between patients’ choices regarding the most important motivational factor and their demographic characteristics in patients with stroke and in those with fracture.

## DISCUSSION

To the best of our knowledge, the present study is the first to investigate the similarities and differences between patients’ and clinicians’ perceptions of the most important factors for motivating patients to engage in rehabilitation. The three motivational factors most frequently endorsed by patients were identical to those endorsed by clinicians. Furthermore, patients had more diverse preferences for motivational factors than clinicians. These findings broaden our understanding of motivation in patients-centered rehabilitation and may provide clinicians with helpful information for effectively motivating patients to engage in rehabilitation.

Participants in this study were recruited from more than 10 hospitals. Moreover, the sample size per group was determined using a priori sample size calculation. Consequently, this study had a large sample size and sufficient response rates (> 60%),^42^ which can potentially reduce nonresponse bias.^38^ Furthermore, we prepared the survey according to existing guidelines and conducted pretesting procedures, such as expert reviews and pilot testing, to establish content and response process validity.^38,39,43^ Therefore, we expected that these procedures would strengthen the reliability and generalizability of our findings.

### Comparison of patients’ and clinicians’ perceptions of important factors for motivating patients to engage in rehabilitation: similarities

The first novel finding regarding similarity in perceptions is that not only clinicians, but also patients consider “goal setting” and “practice related to the patient’s experience and lifestyle” as most important. Previous theoretical, experimental, and observational studies have reported that these two factors are essential components of motivation.^25,29,37,44-47^ Thus, many clinicals use them as the key-motivational strategies in convalescent rehabilitation wards. During rehabilitation, patients may share the intentions of clinicians and recognize them as valuable to overcome difficulties. Our study suggests that these strategies can be the core-motivators supported not only by medical evidence but also subjective perceptions of patients and clinicians. Another novel finding is that “realization of recovery” is considered the most important by both patients and clinicians. “Realization of recovery” would be associated with positive achievement emotions that follows success in rehabilitation.^48^ Several studies indicate that patients’ emotions at the initial stage of rehabilitation predict their subsequent motor performance and recovery after brain injury.^49-51^ However, it remains unclear if success at rehabilitation influences the development of positive emotions and motivation in a real rehabilitation setting. Along with previous qualitative studies,^52,53^ our result suggests that the reverse direction may be effective as a core motivator of rehabilitation, and the relationship between achievement emotions and outcomes might be reciprocal rather than one directional in rehabilitation. This hypothesis should be investigated in future studies.

### Comparison of patients’ and clinicians’ perceptions of factors important for motivating patients to engage in rehabilitation: differences

Of nine motivational factors endorsed by more than 5% of patients as the most important, “medical information” and “control of task difficulty” were preferred more by patients over clinicians. Previous qualitative studies of individuals with physical disabilities have reported that information regarding the benefits of rehabilitation programs is an important factor in increasing patients’ motivation.^6,9,52,54-56^ Additionally, rehabilitation programs that combine exercise therapy with information provision and therapeutic patient education have shown positive outcomes in patients with a range of neurological and orthopedic disorders.^57-64^ Regarding the influence of the level of task difficulty on motivation, an experimental study has reported that participants exert less effort in difficult trials compared to easy trials.^65^ Failure feedback has been reported to undermine learning motivation, because it decreases people’s confidence in their overall ability to pursue their goals and their general expectations of success.^66^

Thus, strategies, such as explaining the rehabilitation process and providing a practice task that is achievable with little effort, may be more effective in motivating patients than clinicians think. Therefore, when determining which motivational strategies to use, clinicians should consider individual patient preferences regarding motivational factors. Patient information, such as demographic characteristics and personality attributes, and the patient’s reactions to a presented motivational strategy, may help clinicians better understand patient preferences.

### Associations between patients’ choices regarding the most important motivational factors and their demographic characteristics

The results of this study indicate that preferences for motivational factors vary depending on the patient’s age and hospital stay length. We found that “goal setting” and “realization of recovery” were preferred by relatively younger patients (under 65 years of age) than older patients. Relatively young patients who were hospitalized in convalescent rehabilitation wards often aim not only to improve their ability to perform activities of daily living but also to engage in social participation, such as returning to work. Therefore, setting goals that are valuable to these patients and helping them experience positive achievement emotions may be especially important for enhancing active participation in rehabilitation.

Additionally, patients with shorter hospital stays were more likely to consider “medical information” to be the most important motivational factor. This result suggests that interventions such as information provision^57-59^ and therapeutic patient education^60-64^ are effective for increasing the motivation of patients in the early period after admission to the convalescent rehabilitation ward. These findings may help clinicians use different motivational strategies tailored to the patients’ conditions.

### Limitations

A primary limitation of this study is that all of the participants were recruited in Japan, potentially limiting the international generalizability of our findings. Nevertheless, the present results support the results of previous studies from different countries.^4-6,9,52,54,55,67-73^ An international survey would improve the external validity of our findings. Another potential limitation is that the opinions of patients with stroke and patients with fracture might have been overstated in the current sample. Similarly, the responses of physical therapists may have been overstated in clinicians’ perceptions. However, patients with stroke and patients with fracture account for approximately 80% of inpatients in convalescent rehabilitation wards in Japan.^35^ Additionally, rehabilitation clinicians working in the convalescent rehabilitation wards consist of 45.7% physical therapists, 28.6% occupational therapists, 14.3% physicians, and 11.4% speech-language-hearing therapists.^35^ Thus, the percentage of patients with stroke, patients with fracture, and physical therapists in our sample may be consistent with the actual situation.

## CONCLUSIONS

The three motivational factors most frequently selected as the most important by patients were identical to those selected by clinicians, suggesting the existence of the core motivational strategies considered by both patients and clinicians. Additionally, patients exhibited more diverse preferences regarding motivational factors compared with clinicians, as there were motivational factors preferred by patients over clinicians. Therefore, clinicians are required to consider individual patient preferences to promote patient-centered care in rehabilitation when determining the most appropriate strategies.

## Data Availability

All data produced in the present study are available upon reasonable request to the authors.

## Author Contributions

Dr Tanaka has full access to all of the data in the study and takes responsibility for the integrity of the data and the accuracy of the data analysis.

*Concept and design:* Oyake, Tanaka.

*Acquisition, analysis, and interpretation of data:* All authors.

*Drafting of the manuscript:* Oyake, Tanaka.

*Critical revision of the manuscript for important intellectual content:* Oyake, Tanaka.

*Statistical analysis:* Oyake.

*Funding:* Tanaka.

*Administrative, technical, or material support:* Yamauchi, Inoue, Sue, Ota, Ikuta, Ema, Ochiai, Hasui, Hirata, Hida, Yamamoto, Kawai, Shiba, Atsumi, Nagafusa.

*Supervision:* Oyake, Yamauchi, Tanaka.

## Additional Contributors

The authors thank Masafumi Sugasawa, Tsuyoshi Tatemoto, Yuto Goto, and Ayumi Mochida at the Tokyo Bay Rehabilitation Hospital for their help and support. They were not compensated for these contributions.

